# Mental Health During COVID-19: A Qualitative Study with Ethnically Diverse Healthcare Workers in the United Kingdom

**DOI:** 10.1101/2021.12.13.21267718

**Authors:** Irtiza Qureshi, Mayuri Gogoi, Amani Al-Oraibi, Fatimah Wobi, Jonathan Chaloner, Laura Gray, Anna L Guyatt, Laura B Nellums, Manish Pareek, On behalf of the UK-REACH Collaborative Group

**Author notes:** Joint senior authors (Laura B Nellums, Manish Pareek).

## Abstract

**Introduction:** Healthcare workers are experiencing deterioration in their mental health due to COVID-19. Ethnic minority populations in the United Kingdom are disproportionately affected by COVID-19, with a higher death rate and poorer physical and mental health outcomes. It is important that healthcare organisations consider the specific context and mental, as well as physical, health needs of an ethnically diverse healthcare workforce in order to better support them during, and after, the COVID-19 pandemic.

**Methods:** We undertook a qualitative work package as part of the United Kingdom Research study into Ethnicity and COVID-19 outcomes among healthcare workers (UK-REACH). As part of the qualitative research, we conducted focus group discussions with healthcare workers between December 2020 and July 2021, and covered topics such as their experiences, fears and concerns, and perceptions about safety and protection, while working during the pandemic. The purposive sample included ancillary health workers, doctors, nurses, midwives and allied health professionals from diverse ethnic backgrounds. We conducted discussions using Microsoft Teams. Recordings were transcribed and thematically analysed.

**Findings:** We carried out 16 focus groups with a total of 61 participants. Several factors were identified which contributed to, and potentially exacerbated, the poor mental health of ethnic minority healthcare workers during this period including anxiety (due to inconsistent protocols and policy); fear (of infection); trauma (due to increased exposure to severe illness and death); guilt (of potentially infecting loved ones); and stress (due to longer working hours and increased workload).

**Conclusion:** COVID-19 has affected the mental health of healthcare workers. We identified a number of factors which may be contributing to a deterioration in mental health across diverse ethnic groups. Healthcare organisations should consider developing strategies to counter the negative impact of these factors. This paper will help employers of healthcare workers and other relevant policy makers better understand the wider implications and potential risks of COVID-19 and assist in developing strategies to safeguard the mental health of these healthcare workers going forward, and reduce ethnic disparities.

**Key messages:** *What is already known about this subject:*

- Healthcare Workers (HCWs) are experiencing deterioration of their mental health due to COVID-19
- Ethnic minority populations and HCWs are disproportionately affected by COVID-19
- More research is needed on the specific factors influencing the mental health of ethnically diverse healthcare workforces

*What are the new findings:* Prominent factors influencing the mental health and emotional wellbeing of this population include: - anxiety (due to inconsistent protocols and policy)
- fear (of infection)
- trauma (due to increased exposure to severe illness and death)
- guilt (of potentially infecting loved ones)
- stress (due to longer working hours and increased workload)

*How might this impact on policy or clinical practice in the foreseeable future:*

- Healthcare organisations should consider the specific circumstances of these staff and develop strategies to counter the negative impact of these factors and help safeguard the mental health of their staff

## INTRODUCTION

Recent evidence suggests that the COVID-19 pandemic is contributing to increased levels of stress, anxiety and trauma for various populations in the United Kingdom (UK) [1]. One of these affected populations is healthcare workers (HCWs) [2], with evidence that previous pandemics have had a significant negative impact on the mental health of this population [3].

In addition, recent research has highlighted that certain groups within society (such as people belonging to ethnic minority backgrounds) have been disproportionately affected by the pandemic. For example, figures from the Office for National Statistics (ONS) show that ethnic minority populations in the UK are experiencing higher death rates and poorer health outcomes than white ethnic groups (ICNARC, 2020). While ethnic minority groups make up 14% of the UK population (Office for National Statistics, 2012) they represent 34% of COVID-19 cases admitted to critical care (ICNARC, 2020). We also know that this overrepresentation applies to both the general population as well as the healthcare workforce [4]. Individuals from ethnic minority backgrounds make up 19.1% of the National Health Service (NHS) workforce in England [5], yet they made up 63% of COVID-19 related healthcare staff deaths in the initial stages of the pandemic [6,7].

It is imperative that healthcare workers are appropriately supported and protected from physical and mental harm during the pandemic, as the World Health Organisation (WHO) have rightly identified them as ‘our most valuable resource for health’ [8]. It will take some time to determine the overall impact of the pandemic, however we need to know more about what factors are contributing to poor mental health and driving disparities within the diverse healthcare workforce. It is important that those employing ethnic minority healthcare workers consider their particular context in order to better protect their physical and mental health, retain them and reduce attrition, and professionally support them during and after the COVID-19 pandemic.

There is increasing quantitative evidence that HCWs are experiencing poor mental health outcomes as a result of the COVID-19 global pandemic. Research with over 2000 HCWs in Italy showed that 63.2% of participants reported COVID-related traumatic experiences at work and 53.8% (95% CI 51.0%–56.6%) showed symptoms of post-traumatic distress; moreover, 50.1% (95% CI 47.9%–52.3%) showed symptoms of clinically relevant anxiety and 26.6% (95% CI 24.7%–28.5%) symptoms of at least moderate depression [9]. A smaller scale study from India showed that of 197 HCWs assessed, a large proportion reported symptoms of depression (92, 47%), anxiety (98, 50%), and low QoL (89, 45%) [10]. Work related stressors were associated with 46% increased risk of combined depression and anxiety (95% CI: 1.15–1.85).

In the UK, there is an emerging evidence base that demonstrates similar experiences and outcomes for HCWs. An online survey of over 1000 UK HCWs during the first wave of the pandemic found nearly 58% of respondents met the threshold for a clinically significant mental disorder (PTSD = 22%; anxiety = 47%; depression = 47%) [11]. Furthermore, drivers behind some of these results seemed to revolve around participant fear of infecting others, access to PPE, and redeployment to areas with high exposure to COVID-19. Our research aims to add to the scarce qualitative evidence base on this subject to provide more in-depth insight into the lived experience of HCWs from diverse ethnic backgrounds

## METHODS

### Study design and setting

We conducted a qualitative sub-study as part of the United Kingdom Research study into Ethnicity and COVID-19 outcomes among healthcare workers (UK-REACH). This sub-study was specifically designed to explore in-depth HCWs’ experiences, their perceptions of risks, fears and concerns, perceptions about safety and protection, support and coping mechanisms available while working during the pandemic, and attitudes and perceptions about the COVID-19 vaccine [2].

### Study population

We included adult (≥16 years of age) HCWs with experience of working in healthcare settings during COVID-19, including both clinical and ancillary staff, in England, Wales, Scotland and Northern Ireland. Purposive sampling was utilised to recruit 164 HCWs from different ethnicities, genders, job roles, migration statuses, and UK regions to obtain a diverse sample. One-to-one interviews and focus group discussions were conducted between November 2020 and July 2021. As part of a rapid response to disseminate urgently needed evidence, this paper reports data from the focus groups conducted as part of the overall study.

### Data collection

Due to existing restrictions on travel and social distancing, the study was conducted remotely, and all processes including, recruitment, consent, and data collection were completed online. Focus groups were conducted through Microsoft Teams. Key topics covered included: participants’ experiences of working during COVID-19; their fears and concerns at work and outside of work; perceived risk factors; challenges faced in accessing information to keep themselves safe; concerns around stigma, discrimination and racism; facilitators and coping mechanisms; and perceptions about COVID-19 vaccination. Following their participation, a gift voucher was given to HCWs in recognition of their contribution to the research. All discussions were recorded with prior permission of the participants, using the recording feature in Microsoft Teams. Recordings were anonymised and transcribed by professional transcribers, and checked for accuracy by the research team (IQ, MG, FW, AAO, LB).

### Analysis

A thematic analysis approach was broadly utilised [12], combining data-driven inductive coding with deductive coding based on *a priori* themes on the mental health of HCWs during pandemics (such as COVID-19) found in the existing literature. IQ led the coding and theme development for this paper. The coding framework was then shared with the team, and developed iteratively through discussion. The final themes were arrived at after no new themes were identified from the iterations, and all the team members were in agreement with the generated codes and themes.

## FINDINGS

### Demographic data

Sixteen focus groups were carried out with a total of 61 participants. Discussions had a duration of about 1.5 hours, with group sizes varying from 2 to 7 members. The details of participants are provided in Table 1. The first column in the table details the variables for participants, the second column shows the total sample for the overall study, and the final column provides the details of the 61 participants who took part in the focus groups that were analysed for this paper.

**Table 1:** Demographic characteristics of participants.

| **Variable** | **Total (N=164)** | **Sample (n=61)** |
| --- | --- | --- |
| **Sex** |  |  |
| Male | 63 | 26 |
| Female | 100 | 35 |
| Other | 1 | 0 |
| **Age, median (IQR)** | **42 (32-53)** | **46 (22-69)** |
| **Ethnicity** |  |  |
| Asian | 65 | 22 |
| Black | 29 | 15 |
| Mixed | 15 | 6 |
| White | 49 | 17 |
| Other | 6 | 1 |
| **Job Role** |  |  |
| Doctors | 44 | 13 |
| Nurses & Midwives | 30 | 12 |
| Allied Health Professionals* | 62 | 15 |
| Ancillary Health Workers | 28 | 21 |
| **UK Region** |  |  |
| England | 140 | 52 |
| Scotland | 7 | 1 |
| Wales | 3 | 0 |
| Northern Ireland | 10 | 5 |
| Unknown | 4 | 3 |

### Contributory factors to poor mental health

Five key themes were identified in the thematic analysis, including: anxiety (due to inconsistent protocols and policy); fear (of infection); trauma (due to increased exposure to severe illness and death); guilt (of potentially infecting loved ones); and stress (due to longer working hours and increased workload) (please see figure 1). Each theme is presented in detail below:

**Figure 1.**
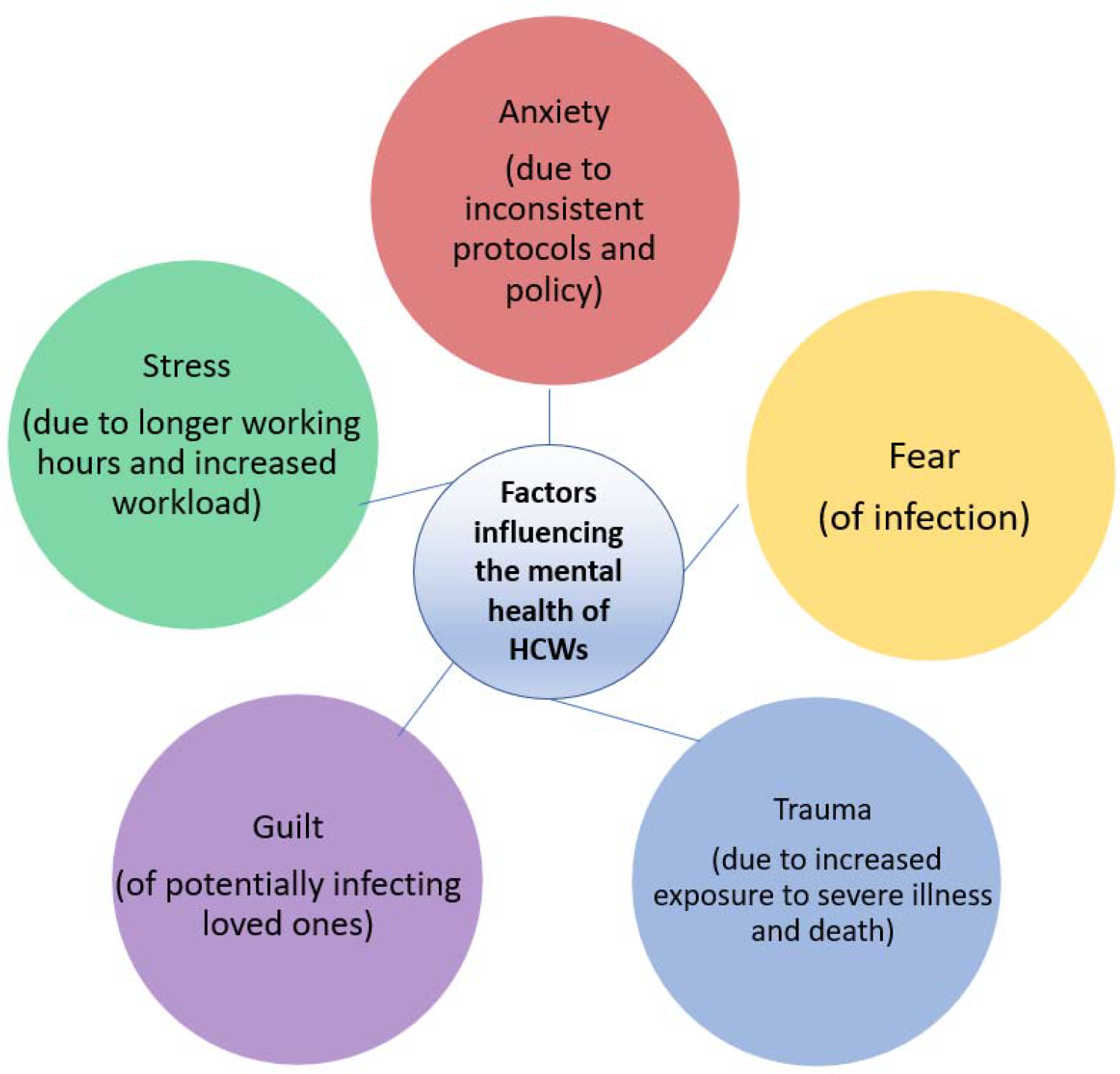
Factors influencing the mental health of Health Care Workers

#### Anxiety (due to inconsistent protocols and policy)

Participants reported feeling an increased level of general anxiety as professional practice guidance fluctuated over days and weeks, early in the pandemic. A clear lack of policy and planning was apparent. This specific confusion around ever-changing guidance for PPE and professional practice added to the general anxiety and confusion in relation to this novel virus and the global pandemic. Participants described that their anxiety levels were raised due to an implicit understanding on the part of the HCWs that employers did not actually know how to appropriately respond to the crisis. Several aspects of practice and guidance were highlighted by participants including infection control:

> *“Anxiety was the first thing. Anxiety and just not sure where we were going and just confusion. The first pandemic, it was just anxiety and confusion and didn’t really know what this meant*.*” (FG12/ F, Doctor)*
>
> *“Personally I was worried, but we’ve always dealt with infections, but this infection was difficult, in there wasn’t clear-cut guidance around, everybody was apprehensive and felt like nobody actually really knew how to handle it at the start anyway*.*”* (*FG 17, M1, Doctor)*
>
> *“It was quite a lot because you know the guidelines were changing almost every – it’s like you get to work today and then the next morning, they’ve come up with something else and when that happens, the way the infection control does things*…*So those are some of the things that are quite – it wasn’t a one-off thing, it was happening frequently*.*” (FG12, M2, Doctor)*
>
> *“I felt like they were still navigating through the policies themselves… it’s the blind leading the blind basically at one point – that’s how it felt*.*” (FG 17, F1, Ancillary Health Worker)*

Other aspects of practice and guidance where inconsistencies and resulting anxiety were described included use of Personal Protective Equipment (PPE) within and across ethnically diverse teams in healthcare settings:

> *“We never worried about getting Covid because we were told if people were passed, they’d died, you couldn’t get it. So we didn’t, apart from wearing normal aprons and gloves and everything which you do anyway, we didn’t wear masks or anything…And then obviously new rules came in where suddenly everybody had to wear masks, even in the mortuary, and it all felt a bit too late, to be honest*.*” (FG2, F2, Ancillary Health Worker)*
>
> *“So at the beginning of the pandemic nobody – so we were office people, no PPE at all, no measures at all. After a couple of months, yeah, probably May/June or something like this then yes, they told us that we have to wear the PPE and the face masks all the time…The following week ‘you have to wear something more’, the following week ‘you have to wear something less’. So it was hard for us to reassure the staff that even with those changes*.*”* (*FG5, F4, Ancillary Health Worker)*
>
> *“It changed all the time, like what you should be wearing and should you wear a visor, shouldn’t you? Should you wear – and people got really anxious about that. One day they were saying ‘no gloves, you have to have your sleeves rolled up’, whereas before we were wearing the gowns and people were just saying ‘I’m just not going to go in then’…it kind of gave everyone a bit of anxiety really because you kind of thought well why are you changing the protocols all the time?*” (*FG13, M2, Allied Health Professional)*

#### Fear (of infection)

Participants were very aware of the heightened risk of infection they were facing in their roles. Ethnic minority staff were acutely aware of the disproportionate impact on them and their communities. They reported bereavements, in personal and professional capacities, and this fed into their personal fears of infection. They also questioned the rationale behind some of the decisions their employers made, which they felt put them at increased risk of exposure to the virus:

> *“We were all kind of anxious about the virus obviously*…*When they called us to help out I knew OK, we were also going to be at the forefront and that meant that I would also be vulnerable to getting the virus. So the anxiety became more evident at that time, yeah*.*” (FG12, M1, Doctor)*
>
> *“There was a personal fear of knowing that my colleagues within even our hub had contracted Covid, again as I said earlier, there was an unseen virus, you didn’t know where it started, you didn’t know where it was going to hit*.*” (FG8, M3, Allied Health Professional)*

#### Trauma (due to increased exposure to severe illness and death)

Although many of the healthcare workers had experienced severe illness and death of patients as part of their work experience, the scale of illness and death and resulting psychological impact of this pandemic was clear in their responses. Again, staff from ethnic minority backgrounds reported a heightened awareness of the illness and death in their communities:

> *“I felt, it was almost like being in a dreamlike state, if that makes sense, like was this happening, as such?*…*I sometimes think oh, I’m going to wake up and this is going to be one big horrible nightmare*.*” (FG2, F2, Ancillary Health Worker)*
>
> *“Everybody who was dying seemed to be BAME [Black, Asian, and Minority Ethnic group] and it was just, I would come in and a patient that I was working on the day before and thought it was going really well and I thought my intervention, you know, I’d made some kind of difference and then I rock up the next day and they’re not there anymore and it just seemed to happen two to three times each day*.*” (FG4, F2, Allied Health Professional)*
>
> *“Every day you leave your house and you’re going to work and it’s more or less like you are in a war zone because you’re trying to protect your life and at the same time you’ve got these very poorly patients that are gasping for relief and so you’re just drawn between two things. For me I would just describe it as if we were in a war zone and every moment is like you’re getting onto the ward and the whole atmosphere is so tense. The emotional, psychological, physical, mental exhaustion, because the time you take to see one patient, normally I would have been able to see five patients*.*” (FG12, M2, Doctor)*

One participant reported being redeployed into a role where they faced exposure to death on a daily basis, having never worked in a clinical capacity before:

> *“So I came from a position of never being clinical, never seeing somebody that had passed away, because I literally, I actually had a fear of dead bodies before I went and worked in the mortuary…Seeing all that death and it was literally I woke up one day and I was like ‘all I see is death. All I see is death. There’s no life*.*’ And I think that was what got me after a while*.*” (FG2, F2, Ancillary Health Worker)*

Some participants reported the cumulative psychological impact of the illness and related bereavement in both professional and personal capacities. The scale as well as personal connection with loss and death seemed to be a clear factor impacting on their mental state. Professional examples included:

> *“I remember the reaction of the second Covid positive when I was about to approach him and he stopped me and he said, ‘Don’t come near me because I have the virus,’ and he was in tears, he was crying*.*” (FG3, F, Nurse)*
>
> *“I had a [teenager] die in front of me because, again, you know, keeping it anonymous but [they] had [a comorbidity] and when you have [it] your impact of the Covid infection is literally a different level…And [they] were well until they weren’t and [they] died very quickly, as in [they] deteriorated very fast*.*” (FG8, M1, Doctor)*

#### Personal loss was also discussed by the participants

> *“I’ve lost 16 friends already with Covid and I don’t know if I’m still counting and I don’t want to count any more because I think it’s just relentless*.*” (FG3, F3, Nurse)*
>
> *“We were doing monthly webinars to begin with, it was all very well-intended and then the person who actually does our social media died two weeks ago of Covid*.*” (FG4, F1, Allied Health Professional)*

#### Guilt (of potentially infecting loved ones)

Many participants spoke of the guilt they felt regarding potentially infecting their family and loved ones. They were very aware of the heighted risk of infection they faced due to their professional role, but felt they were then passing on that risk to those that they lived with. This guilt had a clear impact on their mental wellbeing:

> *“For me it was that guilty feeling, and still is, that I am the one who’s going to kill somebody because I’m going into an area where I know there’s Covid patients and then coming back to people who could actually stay pretty safe, if it wasn’t for me*…*I felt like I was some kind of lethal weapon walking around!” (FG15, F2, Nurse)*
>
> *“At home I have a [young] son who suffers with a [comorbidity], so I have to be very very careful because I have a vulnerable child as well at home that I need to protect*.*” (FG3, F1, Nurse)*
>
> *“If I have to die, let me die with a heart attack rather than Covid positive and I bring that back and infect my children…Not only scared for myself, again, but probably as we all felt, we are more scared for our family than ourselves. I was extremely scared for my husband… now if I have to go somewhere within the hospital, I avoid it. If I don’t have to, I don’t. I really avoid going out. I’m scared. I’m scared*.*” (FG5, F3, Ancillary Health Worker)*
>
> *“I’m then putting my family and friends at risk. I feel like I’m the one that is the danger to everybody now*.*” (FG15, F2, Nurse)*

#### Stress (due to longer working hours and increased workload)

A number of HCWs reported having to work longer hours during this period, describing the increased workload and expectation that they worked beyond their contracted hours:

> *“So my work pattern now should be 9 to 5. Obviously most of us aren’t working those at the moment. So at the moment it’s about 7 to 8*.*” (FG2, F1, Nurse)*
>
> *“You’re supposed to be just working Monday to Friday 8 to 5 or 7 till 4, but sometimes we’ll go in at 7 and at 10 o’clock I’m still there*.*” (FG3, F1, Nurse)*
>
> *“We had two teams working there and I was there from 7 till 11 at night, basically looking after one team and we did everything and I worked 14 days on, 2 days off. It was just – and like weekends, there was no break, there was no break from it*.*” (FG2, F2, Ancillary Health Worker)*

Participants described how they felt that they felt they had an individual responsibility to help cover the increase in demand in patient care, however this came at a cost for them in terms of increased stress, pressure, and fatigue:

> *“I would literally be in tears because I cannot cope with the stress anymore. I cannot cope with they want this, this, this and they want it yesterday. I can’t do it*.*” (FG5, F3, Ancillary Health Worker)*
>
> *“You know, each day of the week we go to work we’re carrying baggage on our backs, a baggage of stress, of anxiety, of saying ‘make sure I do it right*.*’” (FG12, F, Doctor)*

Participants also highlighted the increased workload that each of them faced during the period. Healthcare organisations lacked the capacity to deliver usual levels of care. The increase in work included the diversity of responsibilities HCWs took on, as well as the increase in patients that each of them was expected to care for:

> *“I was doing everything. I was doing management, I was running the ward, I was teaching, because so many people in our ward, they were actually ill or shielding, which is understandable, so they were not around*.*” (FG3, F2, Nurse)*
>
> *“I came back to work after being unwell and then a few days later it was just me with (nearly 30 patients) - 4 of them were (intubated) – because everyone else was unwell*.*” (FG3, F2, Nurse)*

The reporting of chronic understaffing was coupled with the direct impact on HCWs and how many patients they had to care for. The mental strain of this was apparent in their testimonies:

> *“It’s normally one to one nursing but there were some days when it was one to six, one nurse to six patients, with an average of say four or five. So the nurse was just running around. So we were just keeping an eye on the patients, washing them, doing the ventilators, suctioning, that kind of thing, eye care – just all those kind of little bits that I actually found out later are actually a really important part of intensive care*.*” (FG13, M2, Allied Health Professional)*
>
> *“Three or four staff who had to shield because of…different conditions, so it was just very very difficult…Normally we’d have about 280 patients on the caseload and there were 380*.*” (FG6, F1, Doctor)*

## DISCUSSION

In a nationwide study of HCWs, we undertook an in-depth exploration of healthcare worker’s experiences while working during the pandemic, and the impact of this on their mental health. The findings from this research generally align with the emerging evidence base that there is an increased psychological burden on HCWs during the COVID-19 pandemic and this is likely due to the staff reorganization, the working intensity, and the anxiety of being exposed to the virus in the healthcare setting and, in turn, of bringing the infection home [13].Other research also points to increased hours and exposure to COVID-19 patients as factors contributing to poorer mental health among HCWs [14], in line with our findings.

However, the findings from this research also suggest that organisational responses to the pandemic were inconsistent. There was a distinct absence of well-defined and consistent policy in regard to pandemic planning and preparedness. The lack of clear and consistent guidance left HCWs at risk of physical as well as mental harm. Our research shows that HCWs were acutely aware of this increased risk. People with different roles within the healthcare workforce were provided with different guidance and different levels of protection. This included (lack of) access to PPE. Some research suggests that NHS staff were put at risk due to insufficient delivery and provision of appropriate PPE, reporting that by April 2020, only 12% of hospital doctors felt fully protected from the virus at work [15].

There are specific circumstances which relate to the experience of ethnic minority HCWs, which our research highlights. These findings align with other research [16] which identified less access to adequate PPE for ethnic minority HCWs. This situation is associated with decreased trust in employers (as evidenced in this research), which in turn is associated with poorer mental wellbeing for ethnic minority HCWs [14]. Inconsistencies in policy guidance may leave HCWs at increased risk of infection from COVID-19, and combined with a lack of trust in employer, may lead to a fundamental sense of betrayal, which can exacerbate existing marginalisation and inequities [17]. This loss of trust and sense of betrayal has been measured in previous research (e.g. using the moral injury event scale, [18]), and provides further evidence of the detrimental psychological impact on the mental health of ethnic minority HCWs during COVID-19.

Furthermore, the findings in this research highlight a wider potential gap in national policy responses to COVID-19. Historically, a number of government commissioned reports have placed patient safety at the heart of quality of healthcare service provision [19,20]. However it seems that when responding to COVID-19, the government’s primary strategy was to retain the required critical care capacity during surges in demand [21]. This meant that existing problems like the longstanding shortage of nurses and other HCWs in the NHS [22] may have thus negatively impacted on patient safety and staff to patient ratios during COVID-19, as evidenced in our findings. Although the NHS has consistently resisted the imposition of minimum staffing ratios [23] in 2019, guidelines from the Faculty of Intensive Care Medicine (FICM) and Intensive Care Society (ICS) described the levels of care required by critically ill patients in hospital according to their clinical needs. The guidance was clear that ventilated patients must have a registered nurse/patient ratio of a minimum 1:1 to deliver direct care [24]. As the second wave of COVID-19 hit the UK in Winter of 2020, NHS England advised hospitals to temporarily suspend the 1:1 ratio for level 3 critically unwell patients [24]. The findings in this paper show the impact of that decision on ethnic minority HCWs and their mental wellbeing. Participants in our research reported how these national decisions resulted in extremely challenging increases in workload and working hours for these HCWs.

The emerging evidence base suggests that ethnic minority communities are more likely to experience poor health outcomes and higher mortality rates due to COVID-19 [25]. Our findings demonstrate that across diverse ethnic groups, HCWs are experiencing mental distress and trauma due to experiencing significant amount of loss and increased exposure to death at work and bereavement of friends and family. This is likely to exacerbate existing structural and systemic inequities, and disparities in the impact of COVID-19 within the health system. Furthermore, for HCWs from ethnic minority backgrounds, these occupational stressors are likely to be compounded with the wider loss and bereavement they are experiencing at the intersections of their professional and personal communities. Despite consistent evidence that people of minority ethnicity backgrounds are at higher risk of catching COVID-19 and dying from it [6,26], and the increased risk of exposure HCWs from these communities face due to their frontline roles, few solutions have been communicated or actioned [27,28]. The findings in this paper evidence the cumulative effect of these relative risks on the mental health of this population.

## CONCLUSION and RECOMMENDATIONS

This paper highlights some of the prominent factors influencing the mental health and emotional wellbeing of HCWs from diverse ethnic backgrounds during the pandemic (see figure 1). Our findings align with the emerging evidence base, which has highlighted staff reorganisation; working intensity; being exposed to the virus; bringing the infection home; and increased hours and exposure to COVID-19 patients as contributory factors to poorer mental health for HCWs. However, our study also highlights the lack of local and national policy and preparedness; lack of access to PPE; potential decrease in trust and sense of betrayal resulting in higher psychological burden for HCWs. Furthermore, the intersectional impact [29] of belonging to an ethnically diverse workforce leading to higher exposure to death and severe illness, personally and professionally may also contribute to a higher psychological burden. Based on these findings, we recommend that healthcare organisations should consider the specific circumstances of HCWs, and ethnic iniquities among staff, to develop strategies to counter the negative impact of these factors on mental health.

## Data Availability

To access data or samples produced by the UK-REACH study, the working group representative must first submit a request to the Core Management Group by contacting the UK-REACH Project Manager in the first instance. For ancillary studies outside of the core deliverables, the Steering Committee will make final decisions once they have been approved by the Core Management Group. Decisions on granting the access to data/materials will be made within eight weeks. Third party requests from outside the Project will require explicit approval of the Steering Committee once approved by the Core Management Group. Note that should there be significant numbers of requests to access data and/or samples then a separate Data Access Committee will be convened to appraise requests in the first instance.

## DECLARATION OF INTERESTS

MP reports grants from Sanofi, grants and personal fees from Gilead Sciences and personal fees from QIAGEN, outside the submitted work. IQ, MG, FW, IQ, AAO, JC and LBN have no competing interests to declare.

## LIMITATIONS

Due to social distancing measures in place at the time, recruitment strategies and data collection had to be conducted remotely and using online technology. This may have affected participation from certain groups who may be less proficient in use of or have less access to digital technology. The fact that this study collected data in real-time as the situation around the COVID-19 pandemic was unfolding may be seen as a strength in terms of relevance but may also mean that some of the participants’ views may have changed from the time of data collection.

## ETHICS

Ethical approval has been received from the London-Brighton & Sussex Research Ethics Committee of the Health Research Authority (Ref No 20/HRA/4718). All participants gave written informed consent.

## Notes

<u>Funding Statement</u> UK-REACH is supported by a grant (MR/V027549/1) from the MRC-UK Research and Innovation (UKRI) and the Department of Health and Social Care through the National Institute for Health Research (NIHR) rapid response panel to tackle COVID-19. Core funding was also provided by NIHR Biomedical Research Centres. MP is funded by an NIHR Development and Skills Enhancement Award and also acknowledges support from the NIHR Leicester BRC and NIHR ARC East Midlands. LBN is supported by the Academy of Medical Sciences (SBF005/1047). This work is carried out with the support of BREATHE—the Health Data Research Hub for Respiratory Health (MC_PC_19004) funded through the UK Research and Innovation Industrial Strategy Challenge Fund and delivered through Health Data Research UK.

### Funding Statement

UK-REACH is supported by a grant (MR/V027549/1) from the MRC UK Research and Innovation (UKRI) and the Department of Health and Social Care through the National Institute for Health Research (NIHR) rapid response panel to tackle COVID-19. Core funding was also provided by NIHR Biomedical Research Centres. MP is funded by an NIHR Development and Skills Enhancement Award and also acknowledges support from the NIHR Leicester BRC and NIHR ARC East Midlands. LBN is supported by the Academy of Medical Sciences (SBF005 1047). This work is carried out with the support of BREATHE the Health Data Research Hub for Respiratory Health (MCPC19004) funded through the UK Research and Innovation Industrial Strategy Challenge Fund and delivered through Health Data Research UK.

